# QJHong Model for Novel Coronavirus Disease 2019 (COVID-19) in the United States

**DOI:** 10.1101/2023.07.30.23293233

**Authors:** Aanand Mehta, Qi-Jun Hong

## Abstract

We present the methodology of the QJHong model, a machine learning predictive model we built to forecast the COVID-19 daily cases, number of daily deaths, fatality rate, reproductive number, and overall trends in the United States (both national and individual states). We measure the accuracy and compare it to other predictive models. Several forecast analyses consistently demonstrate that the QJHong model outperforms other models submitted to the COVID-19 Forecast Hub with regards to forecasting national data. The Forecast Hub is utilized by the Centers for Disease Control and Prevention (CDC) as a means of disseminating official public communications pertaining to the ongoing pandemic. As such, our model has been identified as a premier performer within this context.

## INTRODUCTION

COVID-19, caused by the SARS-CoV-2 coronavirus, is an infectious disease that has become the most severe pandemic of the twenty-first century. It has resulted in over 670 million cases [1] and more than 6.8 million deaths worldwide, causing a state of international disorder. Although first reported in Wuhan, China in December 2019, the United States has had one of the most severe experiences, with over 100 million cases and over 1 million deaths. Despite efforts to prevent and control the spread of the disease, the emergence of variants [2] that can evade the effects of vaccines [3] has worsened the situation. Hence, it is crucial to comprehend and predict the spread of the disease to enable authorities to take necessary precautions and develop solutions more rapidly. Several models [4], including compartmental models such as the SIR (Susceptible-Infected-Removed) [5, 6] and its variations, most notably the SEIR (Susceptible-Exposed-Infectious-Removed) [7, 8] models, machine learning models, and ensemble models [9], have been developed to forecast the progression of the disease.

The SIR model is a widely used epidemiological model that divides a population into three groups: susceptible, infected, and removed. It was developed by Kermack and McKendrick (1991) [10] and represented as N = S + I + R, where N is the total population. The model uses differential equations to calculate the changes in each group over time, with the constants representing the probability of transmission and recovery rate [11], respectively. Before the virus affects a population, all groups are assumed to be empty except for the susceptible group. Two COVID-19 SIR models, BPaganoRtDriven [6] and IowaStateLW-STEM [5], are commonly used for forecasting. The SEIR model, a variation of the SIR model, introduces an Exposed group, E(t), representing individuals who are infected but not yet contagious. This allows for more specific parameter descriptions. Two recognized COVID-19 SEIR models are CU-select [8] and UCLA-SuEIR [7]. The ensemble model aggregates multiple models (e.g. SIR, SEIR, machine learning) to provide a combined forecast, consistently outperforming individual models for various diseases. The COVIDhub-ensemble model [12] is an example of this, incorporating forecasts from over 25 academic, private, and government-affiliated groups, as well as the CDC. The model collects death-predicting forecasts from different models, aggregates them, and evaluates their error with mean absolute error. For inclusion, an individual model must submit a valid 4-week forecast and is equally weighted with other eligible forecasts [13].

Here, we propose a “Mobility-Data model,” the QJHong model we built to forecast COVID-19 daily cases and deaths in the United States. The model is based on machine learning between COVID-19 cases and key predictive attributes, such as mobility data in Google’s Community Mobility Reports. Anonymously tracking the location history of Google users, it compares movement by country to a baseline (set before the pandemic, the median day value from January 3 to February 6, 2020). Further, it takes into account the location history of Google users in often-populated city locations (parks, grocery stores, transit stations). This comprehensive data [15] also takes into account the reported data from the pandemic’s early stages (February 15, 2020) until October 15, 2022. Other attributes include weather, vaccination, and status of the pandemic, which we use to represent people’s awareness and response to the virus. For the calculation of daily deaths, we rely on analytical solutions, i.e., statistical distributions and differential equations, to study the relation between cases and deaths, as we find this approach more reliable and accurate than machine learning.

## METHODOLOGY

### Forecasting new cases: machine learning

In this study, we have developed a machine learning model to calculate the reproductive number R, which enables us to predict future daily new cases. Our model incorporates various key predictive attributes, including mobility data, weather, vaccination rates, and recent COVID-19 status, as well as recurrent features such as the reproductive numbers from 60, 30, and 15 days ago, and location for state-level modeling. We utilize the XGBoost method to learn the relationship between our features and targets.

We have found that predicting future cases is a much greater challenge compared to forecasting deaths, as the pandemic is constantly evolving, and new attributes emerge as the dominant attribute changes. Since the beginning of the pandemic, key factors have shifted from mobility during lockdown to temperature and weather, and from variant types such as alpha, delta, and omicron to vaccination rates. As a result, relying solely on past data to make predictions for the future may not be reliable. Our approach is to (1) accurately predict new deaths from cases, as described in the next section, and (2) forecast new cases as best as possible by incorporating the most important features to capture the key dominating attributes.

### Forecasting new deaths: analytical fitting

To calculate daily deaths, we rely on analytical solutions such as statistical distributions and differential equations to study the mathematical relationship between cases and deaths. Our approach is more reliable and accurate than machine learning, as we have found a clear and simple mathematical relation between cases and deaths.

The Poisson process is utilized to describe the transition from cases to deaths. Let *N* (*t*) denote the new daily cases on day *t, D*(*t*) the corresponding fatality on the same day, and *γ* the fatality rate. While it may appear that *D* equals *N* mulitplied by *γ*, deaths do not occur instantaneously and concurrently with cases. Rather, they conform to the Poisson process, where the number of deaths on day *t* is given by

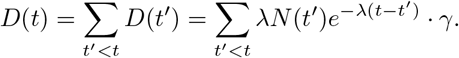

Consider *N* (*t*^*′*^)*· γ* as the number of deaths that will eventually occur as a result of cases from day *t*^*′*^, but which are statistically distributed over time according to the Poisson distribution with decay parameter *λ*. In other words, the daily count of deaths on day *t* is obtained by summing up small counts from all preceding days *t*^*′*^ *< t*, where the deaths are inevitable due to the fatality rate *N* (*t*^*′*^)*· γ*, but the actual death occured on day *t* following a Poisson distribution *λe*^*−λ*(*t−t′*^). This presents the statistical count of individuals who are expected to die on day *t*.

To predict future death counts, we conduct parameter fitting for *λ* and *γ* based on past relationships between *D* and *N*. Using these two parameters, along with a projection of future cases *N*, we can make forecasts for future deaths *D*. To account for the variation of fatality rate over time, we make the assumption that the fatality rate follows a basic exponential relationship in the short term. This enables us to project into the near future and make predictions for fatality.

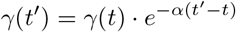

## RESULTS AND DISCUSSION

### Results

As exhibited on Figure 2, our fatality data seven-day average forecast (black curve) well-aligns with the previous forecasts (pale red). However, it is necessary to perform a logical walkthrough of the timelines at which COVID-19 cases, deaths, and fatality rates align with our forecast. This is facilitated by the “times of prevalence” of the Alpha, Beta, Delta, and Omicron variants. In other words, our model precisely depicts the trends of each variant, and even accounts for real-time implementations of vaccine shots, boosters, and historical COVID-19 quarantine restrictions.

We compiled a plethora of COVID-19 databases, such as the CDC COVID-19 timeline, the GISAID Tracking of Variants tool, and several other publications through online search. It must also be taken into consideration that the timeframes mentioned below are rough estimates of the “times of prevalence.” In other words, there is not an objectively defined time when a specific variant is overtaken by another variant; there do exist overlays between multiple variants at the same time (as shown in Figure 1). Thus, the boundaries listed below are not rigid.

**FIG. 1.**
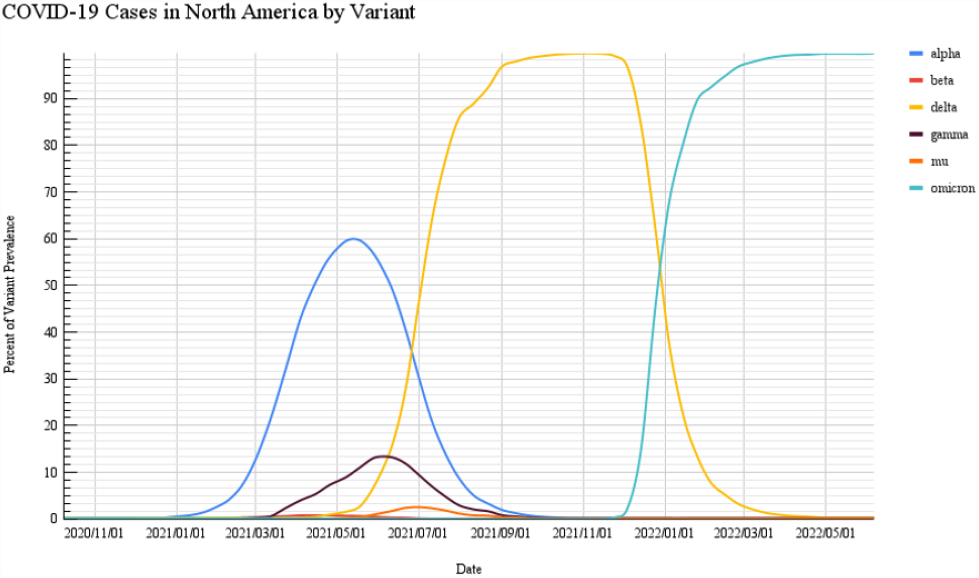
Classifying COVID-19 Cases in North America by Variant: displayed above is a diagram (data sourced from GISAID [14]) showing the percent of variant prevalence over time. Note that the time “2020/11/01 to 2021/01/01” is the time that no variants existed in North America, and all cases consisted of the wild-type strain. In fact, the wild-type strain was widely prominent until 2021/07/01, with the plurality of the “alpha,” “delta,” “gamma,” and “mu” strains.

**FIG. 2.**
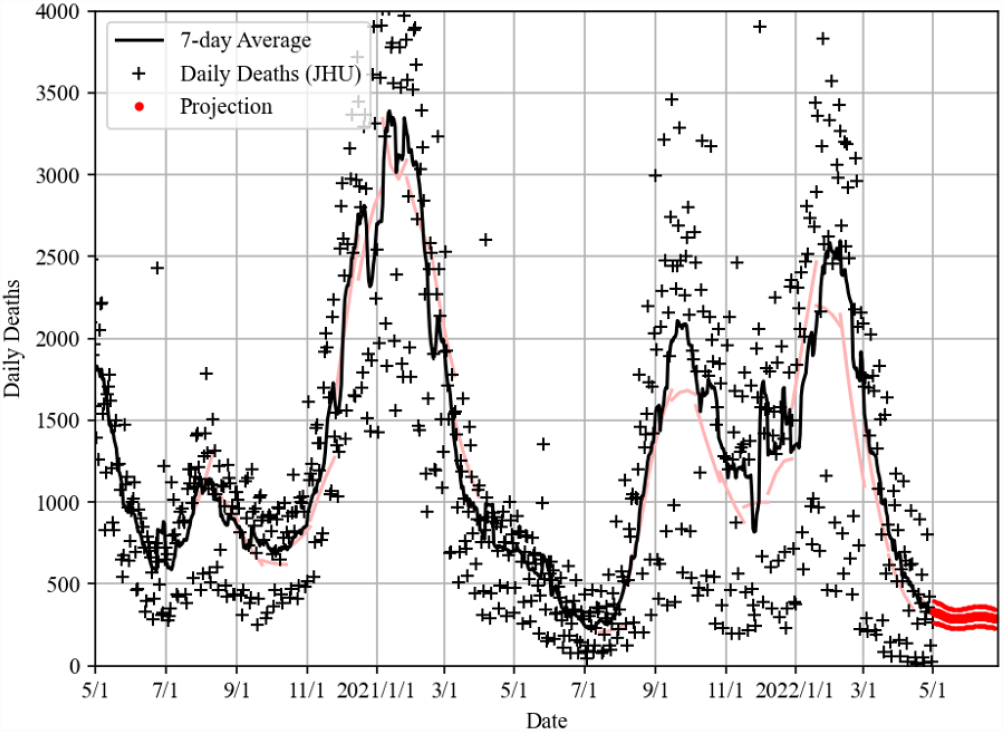
Forecasts generated by our model for the US national fatality rates. The latest forecast is depicted in red, while previous forecasts are displayed in pale red. The actual fatality data, represented by black ”+” symbols, and the corresponding seven-day average, represented by the black curve, are also shown. It is worth noting that the previous forecasts align closely with the actual fatality data.

**FIG. 3.**
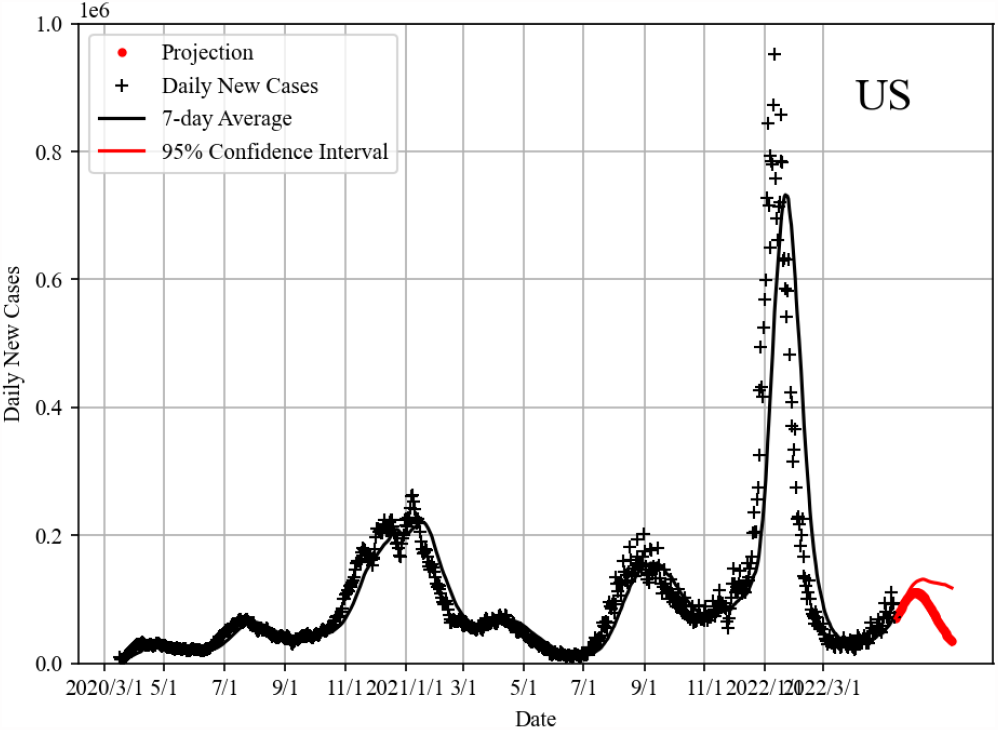
Forecasts generated by our model for the US national case rates. The latest forecast is depicted in red. The actual case data, represented by black “+” symbols, and the corresponding seven-day average, represented by the black curve, are also shown. It is worth noting that the previous forecasts align closely with the actual case data.

**FIG. 4.**
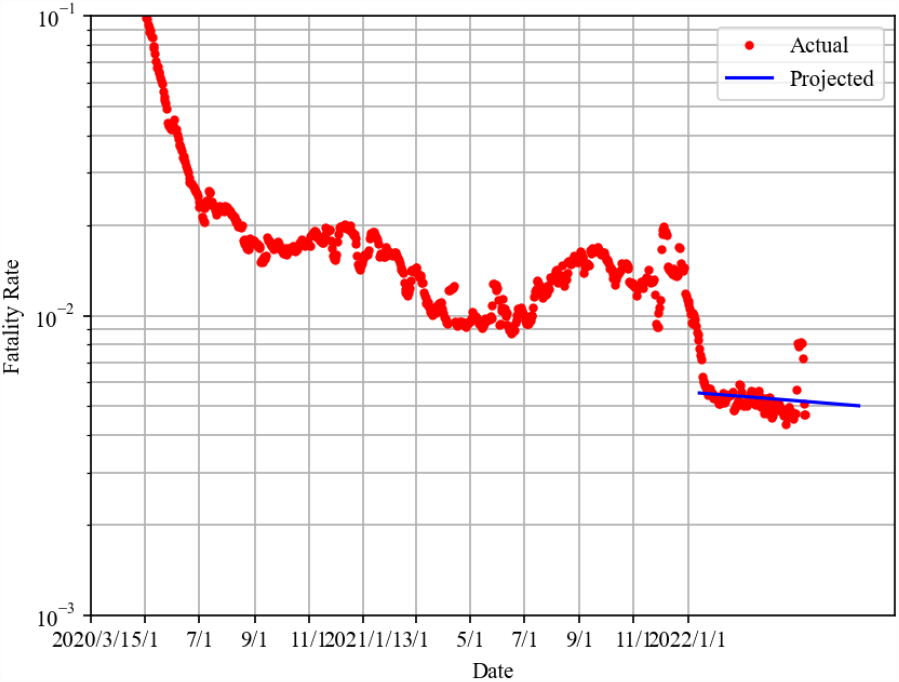
Forecasts generated by our model for the US fatality rates. This forecast utilizes the Poisson process to forecast these rates. The latest forecast is depicted in blue. The actual fatality rate data, represented by red “•” symbols is also shown. It is worth noting that the previous forecasts align closely with the actual case data.

First, the Alpha (B.1.1.7) and Beta (B.1.351) variants arose in the US in December of 2020 and January of 2021 [18], respectively. Here, we are making the decision to group the Beta variant with the Alpha variant, for two reasons. First, for the proximity of times at which both variants were discerned in the US, as aforementioned above, and second, because the Beta variant was quite rare in the US, only having 3,155 total cases in the US, and at its peak (week of 4/19/2021), only consisting of 0.8% of all North American COVID-19 cases [14]. We are characterizing this time period from March of 2021 to June of 2021. Our model displays a spike in daily cases and deaths, but a decreased fatality rate. This is consistent with real-life data, as the Alpha and Beta variants are indeed more fatal and transmissible than the original, wild-type strain. In fact, Washington et al. [19] report that the Alpha variant was 40 to 50 percent more transmissible than the original strain in the United States; Pearson et al. [20] report, assuming that natural immunity provides full protection against the virus, that the Beta variant is 50 percent more transmissible than the original strain. Building off the estimate of the Beta variant’s transmissibility made by Pearson et al., it is safe to assume, in a non-ideal world, that the variant is, in fact, even more contagious. However, beginning in December of 2020, the COVID-19 vaccines were also being introduced to the United States, which is acknowledged by the downtrend in all three statistics from early 2021 until June of 2021. A study published by Al-Aly et al. proved that vaccinations lower the risk of death from COVID19 by 34% [21]. Furthermore, 44% of the US population was fully vaccinated by June 15, 2021 [22] (date chosen because it is near the end of this “time of prevalence”). For this reason, both the fatality rate and daily new cases were at their lowest level in 2021, and daily deaths were quite close to their lowest level in 2021 around June 15, 2021. Thus, it is corroborated that COVID-19 vaccinations play a vital role in preventing the deleterious effects of the disease. Next, the Delta (B.1.617.2) variant arrived in the US in July of 2021 [18]. We are characterizing this time period from July of 2021 to December of 2021. This variant is represented by the spike in daily cases and deaths, and a slight increase in fatality rate. This is logical because the Delta variant has been declared the more severe variant; in fact, the fatality rate should have severely increased, but COVID-19 vaccine booster shots were being introduced in the latter half of this “time of prevalence,” which retaliated against a more drastic increase in fatality rate [23]. Kuehn [24] finds that after being vaccinated with the Johnson & Johnson vaccine and an mRNA-based booster shot, the former is 78% effective in protecting against COVID-19 hospitalizations. Further, 90% protection against COVID-19 hospitalization is established after a total of 3 mRNA vaccinations. It is essential to define that ”Fully vaccinated” denotes receiving both doses of two-dose vaccinations, such as Pfizer-BioNTech or Moderna, or the sole dose of one-dose vaccinations, such as Johnson & Johnson.

Finally, the Omicron (B.1.1.529) variant arrived in the US in November of 2021 [18]. We are characterizing this time period from January of 2022 to present-day. This variant is characteristic of increased transmissibility, yet a lower rate of death. Our model is consistent with this information, as it displays a spike in daily cases and deaths, yet a decreased fatality rate. However, the Omicron variant has been proclaimed as a less deadly variant; this is consistent with our model, as it displays a decreased fatality rate. However, it must be noted that there is a decreasing susceptibility to the virus in early 2022 due to increased seroprevalence within the population. It must be noted, regarding our model, the QJHong Encounter model, that because it is a machine-learning model that relies on past, inputted data, that an increased sample size of the amount of days that a specific variant has been of prevalence increases the future predictability of the virus’ trends. This is because the model can become more accustomed to the specific variant’s characteristics, such as transmissibility or fatality rate.

### Evaluations

The model in question has garnered a reputation for being among the most effective models for predicting fatalities due to COVID-19 in the United States. This paper provides a summary of three separate evaluations conducted by independent sources, namely YouYang Gu’s YYG, Steve McConnell’s CovidComplete, and Carnegie Mellon University’s DELPHI. Figures 5-7 present evidence that our model consistently outperforms a collection of models included in the COVID19 Forecast Hub project. As illustrated in Figure 5, the evaluation conducted by YYG revealed that our model exhibited the highest level of effectiveness among all models evaluated, based on the ranking metric. Additionally, our model performed within the top three models in terms of errors. Figures 6 and 7 indicate that, as per the weekly model scores evaluated by Steve McConnell using COVIDComplete’s ranking system, which awards the highest scores of 100% to the best-performing models, our model was ranked as the second-best model. According to the evaluations conducted by DELPHI, our model demonstrated exceptional long-term accuracy and often outperformed the ensemble model, which is considered one of the best models and widely recognized as the benchmark model.

**FIG. 5.**
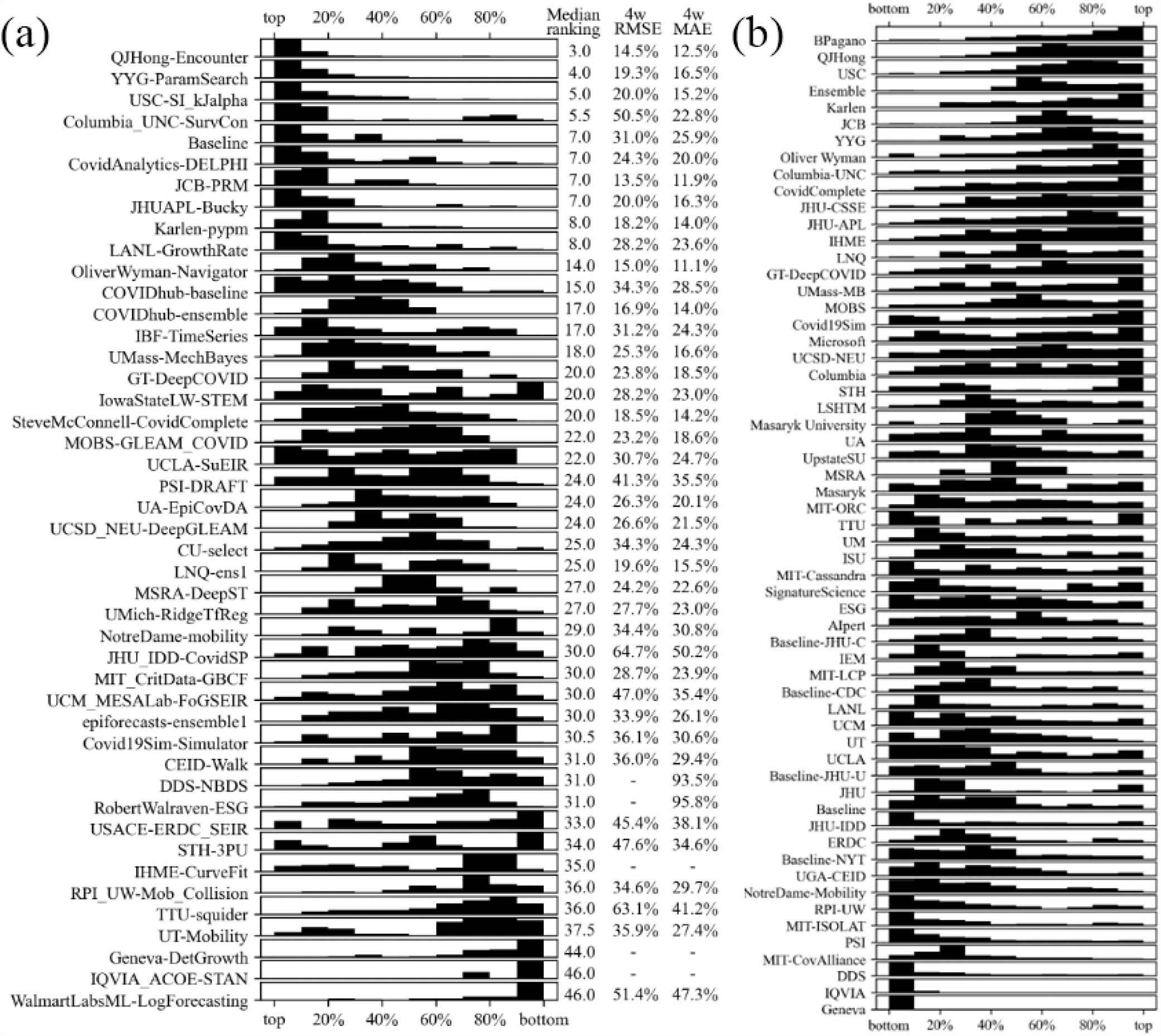
Evaluation results of You-Yang Gu’s YYG [16] and Steve McConnell’s COVIDComplete [17] on the weekly US national fatality rates. Panel (a) depicts the weekly ranking distributions of the top percentile models collected by the COVID19 Forecast Hub, along with median weekly rankings, root mean square errors, and mean absolute errors of the four-week forward forecast. Our model was determined to be the best model in terms of ranking and one of the top three models in terms of errors. Panel (b) showcases the weekly model scores evaluated by Steve McConnell, where the highest scores (100%) were awarded to the top-performing models. Our model was ranked second in this evaluation.

**FIG. 6.**
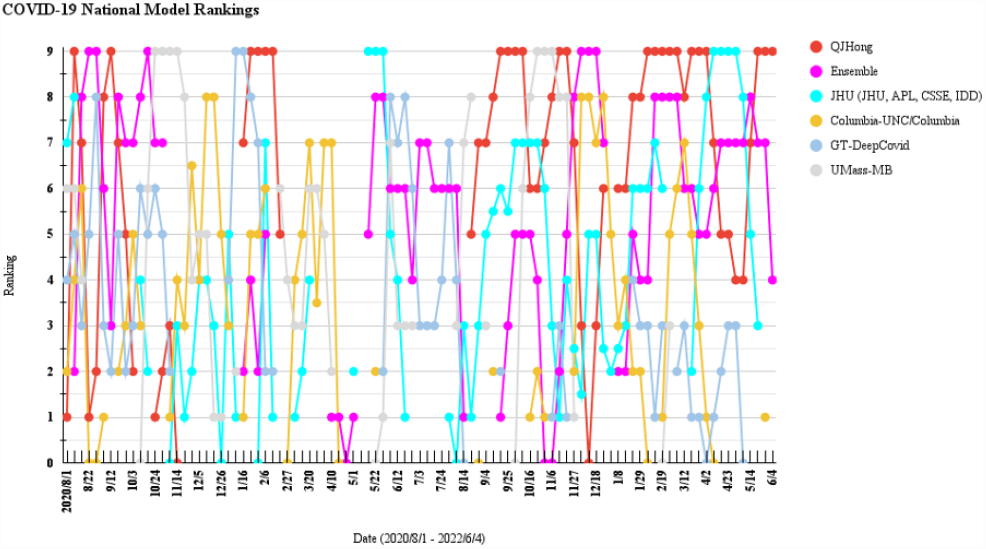
Graphical representation of Steve McConnell’s Forecast Evaluation Rankings [17], comparing our model, QJHong (depicted in red), with five other models (Ensemble, Johns Hopkins, Columbia, Georgia Tech, and University of Massachusetts-Amherst) for predicting COVID-19 cases on a weekly basis. The evaluation spanned from August 1, 2020, to June 4, 2022.

**FIG. 7.**
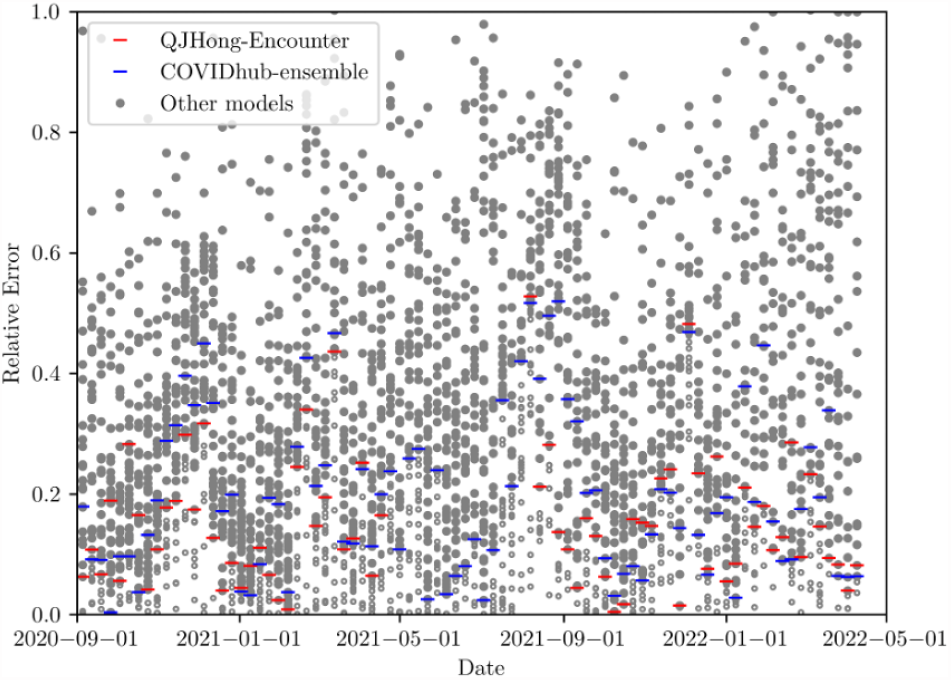
Results of the evaluation conducted by the DELPHI program [25] at Carnegie Mellon University, which assessed the 4-week forward absolute errors of various models on a weekly basis. The performance of most models was found to be volatile, making it challenging for models to consistently achieve top performance. However, over a prolonged period of analysis, QJHong and Ensemble emerged as two of the leading models, consistently outperforming most models (represented by filled gray dots) while only a few models occasionally performed better (represented by open gray circles). It should be noted that although some models were more accurate in certain weekly evaluations, our model demonstrated superior long-term accuracy when evaluated against the vast majority of models.

## CONCLUSIONS

In conclusion, we have presented the methodology of the QJHong model, a machine learning predictive model developed to forecast COVID-19 trends in the United States. Through rigorous evaluation and comparison with other models, our analyses have consistently shown that our model outperforms other models submitted to the COVID-19 Forecast Hub in terms of forecasting national data. It is worth noting that the Forecast Hub is a key platform utilized by the Centers for Disease Control and Prevention (CDC) to disseminate official public communications related to the ongoing pandemic. Hence, our model has been identified as a premier performer within this context, indicating its potential usefulness in guiding decision-making and resource allocation in the fight against COVID-19.

## Data Availability

All data produced in the present study are available upon reasonable request to the authors

## Notes

### Competing Interest Statement

The authors have declared no competing interest.

### Funding Statement

This study was funded by the start-up funding provided by the School of Engineering for Matter, Transport, and Energy, Arizona State University.

